# In-Hospital Mortality in Chagas vs Non-Chagas Heart Failure: A Nationwide Real-World Analysis From the Brazilian Public Health System

**DOI:** 10.64898/2026.04.26.26351771

**Authors:** Camila Nicolela Geraldo Martins, Adriana Aparecida Bau, Guilherme Cordeiro, José R. Matos-Souza, Wilson Nadruz, Andrei Sposito, Carlos Eduardo Rochitte, Ahmad Masri, Michael Jerosch-Herold, Otávio Rizzi Coelho-Filho

## Abstract

**Background:** Chagas cardiomyopathy remains a major cause of heart failure (HF) in endemic regions and is increasingly recognized globally, yet data on in-hospital outcomes are limited. Objective: To assess whether Chagas disease is associated with higher in-hospital mortality among patients hospitalized with HF.

**Methods:** We analyzed a nationwide administrative database from the Brazilian Unified Health System (DATASUS/SIHSUS), including adults hospitalized with HF between April 2017 and August 2021. HF was identified using ICD-10 code I50.x and Chagas disease using B57.x. The primary outcome was in-hospital mortality, evaluated using multivariable Cox models. Results: Among 910,128 HF hospitalizations, 1,082 (0.12%) were associated with Chagas disease. Patients with Chagas were younger but had a more complex clinical profile and higher resource use. In-hospital mortality was higher in the Chagas group (25% vs 12%; p<0.001). After adjustment, Chagas disease remained independently associated with mortality (HR 1.54; 95% CI 1.35–1.75; p<0.001).

**Conclusions:** In this large real-world cohort, Chagas disease was associated with higher in-hospital mortality and greater healthcare utilization. These findings reinforce the high-risk nature of Chagas cardiomyopathy and point to the need for more targeted treatment strategies.

**⍰ What is the clinical question being addressed?:** Chagas cardiomyopathy is a major cause of heart failure in endemic regions and an emerging global health problem, yet real-world data on in-hospital outcomes remain limited. Is Chagas disease associated with higher in-hospital mortality?

**⍰ What is the main finding?:** Chagas disease was independently associated with a 54% higher risk of in-hospital mortality in a large real-world cohort.

---

Chagas disease remains a major cause of heart failure (HF) in endemic regions and is increasingly seen in non-endemic countries due to migration, affecting ∼6–8 million people worldwide, including ∼300,000 in the United States and ∼50,000 in Europe(1). Chronic *Trypanosoma cruzi* infection leads to a distinct cardiomyopathy characterized by progressive myocardial injury, ventricular dysfunction, arrhythmias, and thromboembolic complications(2). Among HF etiologies, Chagas cardiomyopathy has consistently been associated with worse outcomes. A systematic review and meta-analysis including 37 studies and 17,949 patients demonstrated a significantly higher risk of mortality compared with non-Chagas cardiomyopathies (HR 2.26; 95% CI 1.65–3.10)(3). Observational cohorts have reported similar findings(4), even in the context of contemporary HF therapy. In a randomized clinical trial evaluating sacubitril/valsartan in patients with Chagas cardiomyopathy, clinical benefit was observed; however, event rates remained high(5). Despite consistent evidence for worse long-term outcomes, data focused on the in-hospital phase remain limited. A recent analysis from the United States reported higher healthcare utilization among patients with Chagas disease but no difference in in-hospital mortality compared with other HF etiologies(6). We analyzed a nationwide administrative database from the Brazilian Unified Health System (DATASUS/SIHSUS) to evaluate the association between Chagas disease and in-hospital mortality in patients hospitalized with HF. This retrospective, records-based study included adults (≥18 years) admitted between April 2017 and August 2021. HF hospitalizations were identified using ICD-10 code I50.x, with Chagas disease defined by code B57.x.The primary outcome was in-hospital mortality, assessed using multivariable Cox models. Follow-up was defined from admission to discharge or death. The study was approved by the Institutional Review Board of the University of Campinas (CEP-No. 117/2022), with waiver of informed consent.

A total of 910,128 HF hospitalizations were included, of which 1,082 (0.12%) were associated with Chagas disease. Patients with Chagas disease were younger (median 67 [57– 76] vs 69 [59–79] years; p<0.001), with a similar proportion of women (49% vs 48%; p=0.849). The clinical profile differed substantially. Patients with Chagas disease had higher prevalence of hypertension (25% vs 4.4%; p<0.001), diabetes (10% vs 1.9%; p<0.001), atrial fibrillation (5.5% vs 0.7%; p<0.001), prior stroke (2.1% vs 0.3%; p<0.001), renal insufficiency (11% vs 1.6%; p<0.001), and obstructive pulmonary disease (4.6% vs 1.0%; p<0.001).

Hospitalizations were more complex, with longer length of stay (median 7 [4–13] vs 5 [3–9] days; p<0.001) and higher use of renal replacement therapy (3.8% vs 2.2%; p<0.001). Resource utilization was also higher, with greater total hospitalization costs (BRL 1,161 [824–2,783] vs 783 [715–1,254]; p<0.001) and higher cost per day (BRL 225 [122–450] vs 202 [117–358]; p<0.001). In-hospital mortality was higher in patients with Chagas disease (25% vs 12%; p<0.001). Kaplan–Meier curves (Figure 1A) showed early and sustained separation between groups, consistent with higher mortality risk in patients with Chagas disease. In univariable Cox analysis, mortality was associated with older age (HR 1.03; 95% CI 1.03–1.03; p<0.001), female sex (HR 1.12; 95% CI 1.10–1.13; p<0.001), renal insufficiency (HR 1.50; 95% CI 1.46–1.55; p<0.001), ischemic heart disease (HR 1.42; 95% CI 1.35–1.49; p<0.001), obstructive pulmonary disease (HR 1.20; 95% CI 1.14–1.26; p<0.001), and prior stroke (HR 1.79; 95% CI 1.66–1.92; p<0.001). Chagas disease was also associated with mortality (HR 1.53; 95% CI 1.36–1.73; p<0.001). After multivariable adjustment, Chagas disease remained independently associated with in-hospital mortality (HR 1.54; 95% CI 1.35–1.75; p<0.001, Figure 1B).

**Figure 1AB:**
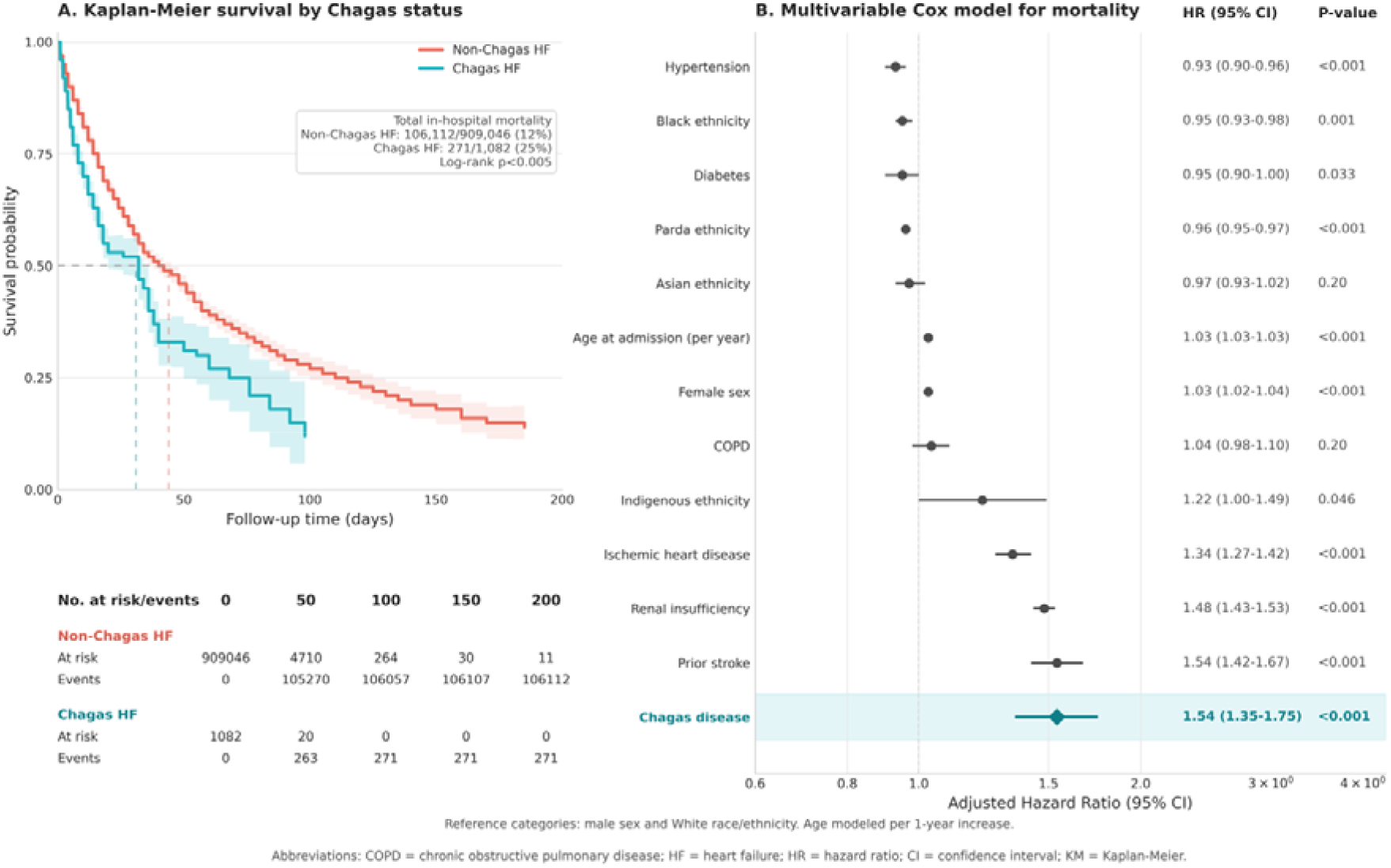
(A) Kaplan–Meier curves for in-hospital survival stratified by Chagas status. Patients with Chagas disease showed significantly lower survival compared with non-Chagas heart failure (HF) (25% vs 12% in-hospital mortality; log-rank p<0.005). Shaded areas represent 95% confidence intervals. (B) Multivariable Cox proportional hazards model for in-hospital mortality. Chagas disease was independently associated with higher mortality risk after adjustment for clinical covariates (adjusted HR 1.54; 95% CI 1.35–1.75; p<0.001). Abbreviations: CI = confidence interval; COPD = chronic obstructive pulmonary disease; HF = heart failure; HR = hazard ratio; KM = Kaplan–Meier.

In this nationwide real-world cohort, Chagas disease was independently associated with higher in-hospital mortality among patients hospitalized with HF. This finding extends prior evidence by showing that the excess risk described in longitudinal studies is already evident during the index hospitalization, suggesting a combination of advanced disease at presentation and increased vulnerability during acute decompensation. The magnitude of association observed here (HR 1.54) is lower than that reported in the meta-analysis (HR 2.26) (3), which is expected given the shorter time horizon of in-hospital outcomes. Our findings also help place results from non-endemic settings into perspective. In the United States, no difference in in-hospital mortality was observed(6). However, that analysis included a relatively very small number of patients with Chagas disease (n=154), representing only a minimal proportion of HF admissions, which substantially limits the ability to detect meaningful differences in mortality(6). Importantly, despite the absence of a mortality signal, that study reported longer hospitalizations and higher healthcare costs among patients with Chagas disease, a pattern consistent with our findings. This apparent dissociation—greater resource utilization without a corresponding increase in mortality— likely reflects limited statistical power rather than true equivalence in risk. In addition, cohorts from non-endemic settings may not fully capture the clinical spectrum of Chagas cardiomyopathy, as patients are often diagnosed earlier or managed within healthcare systems with different access and referral patterns. In contrast, patients in the present cohort exhibited a higher burden of comorbidities, longer hospitalizations, and greater healthcare utilization, all consistent with more advanced disease at presentation and supporting the observed increase in in-hospital mortality. From a broader perspective, Chagas disease is no longer confined to endemic regions. A substantial number of affected individuals now live in the United States and Europe(1), making the condition an increasingly relevant global health problem. From a therapeutic standpoint, the results are particularly informative. Even in the era of contemporary HF therapy, including sacubitril/valsartan, event rates remain high in patients with Chagas cardiomyopathy(5). This persistent risk highlights a key limitation of current management strategies: while guideline-directed therapies are beneficial, they do not fully address the disease-specific mechanisms driving myocardial injury. Chronic inflammation, diffuse fibrosis, microvascular dysfunction, and autonomic imbalance likely remain central to disease progression and clinical instability. The consistently high event rates seen across studies highlight a clear gap in care for patients with Chagas cardiomyopathy. Standard HF therapies may not be enough for this population, and what was once viewed as a regional disease is now becoming a global problem with ongoing migration

This study has limitations inherent to administrative data, including potential coding inaccuracies, underreporting of comorbidities, and limited clinical detail, including HF phenotype. However, in-hospital mortality and HF and Chagas diagnoses were reliably captured, including timing of hospitalization and death. Although the nationwide dataset enhances representativeness, residual confounding cannot be excluded.

In conclusion, Chagas disease was associated with worse in-hospital outcomes and greater healthcare utilization among patients hospitalized with HF. Taken together with prior evidence, these findings reinforce the high-risk nature of Chagas cardiomyopathy and highlight a persistent gap between current treatment strategies and disease-specific pathophysiology. Addressing this gap will require earlier recognition, improved risk stratification, and the development of targeted therapies aimed at the underlying mechanisms of myocardial injury in Chagas disease.

## Data Availability

The data that support the findings of this study are derived from the Brazilian Unified Health System (DATASUS/SIHSUS, https://datasus.saude.gov.br), which is publicly available. Processed data and analytic code may be made available upon reasonable request to the corresponding author.

